# On the impacts of the COVID-19 pandemic on mortality: Lost years or lost days?

**DOI:** 10.1101/2022.06.23.22276812

**Authors:** Valentin Rousson, Isabella Locatelli

**Affiliations:** Center for Primary Care and Public Health (Unisanté), Route de Berne 113, 1010 Lausanne, University of Lausanne, Switzerland

**Keywords:** All-cause mortality, COVID-19, Life expectancy loss, Population life loss, Remaining life expectancy, Years lost

## Abstract

**Objective:** To quantify the (direct and indirect) impacts of the COVID-19 pandemic on mortality for actual populations of persons living in 12 European countries in 2020.

**Method:** Based on demographic and mortality data, as well as remaining life expectancies found in the Human Mortality Database, we calculated a “population life lost” in 2020 for men and women living in Belgium, Croatia, Denmark, Finland, Hungary, Lithuania, Luxembourg, Norway, Portugal, Spain, Sweden and Switzerland. This quantity was obtained by dividing the total number of years lost in 2020 (estimated from all-cause mortality data and attributed directly or indirectly to COVID-19) by the size of the population.

**Results:** A significant population life loss was found in 8 countries in 2020, with men losing an average of 8.7, 5.0, 4.4, 4.0, 3.7, 3.4, 3.1 and 2.7 days in Lithuania, Spain, Belgium, Hungary, Croatia, Portugal, Switzerland and Sweden, respectively. For women, this loss was 5.5, 4.3, 3.7, 3.7, 3.1, 2.4, 1.6 and 1.4 days, respectively. No significant losses were found in Finland, Luxembourg, Denmark and Norway. Life loss was highly dependent on age, reaching 40 days at the age of 90 in some countries, while only a few significant losses occurred under the age of 60. Even in countries with a significant population life loss in 2020, it was on average about 30 times lower than in 1918, at the time of the Spanish flu.

**Conclusions:** Our results based on the concept of population life loss were consistent with those based on the classical concept of life expectancy, confirming the significant impact of COVID-19 on mortality in 8 European countries in 2020. However, while life expectancy losses were typically counted in months or years, population life losses could be counted in days, a potentially useful piece of information from a public health perspective.

## 1. Introduction

As of June 2022, the COVID-19 pandemic that began in 2019 in China have officially killed over 6 million people in the world and this statistic might be underestimated by a factor 3 due to unreliable diagnosis or reporting [1]. Moreover, the COVID-19 pandemic might not only have direct but also indirect impacts on mortality, for example because of delayed medical interventions due to hospital overcrowding [2]. A more accurate indicator to assess the real impact of a pandemic on mortality would be based on all-cause mortality rather than specific (in our case COVID-19) mortality [3]. An estimate of mortality due to COVID-19 can then be obtained by comparing observed all-cause mortality during the pandemic and pre-pandemic years, or with an expected mortality taking into account secular trends in mortality decline.

There are however many ways to summarize mortality in a given year, including the two classic indicators of standardized mortality rate (SMR, sometimes simply called “mortality”) and life expectancy (at birth), which can give quite different results [4]. For example, Locatelli and Rousson [5-6] calculated a 9.2% increase of SMR in Switzerland in 2020 compared to 2019, which corresponded to a decrease of (only) 0.8% in life expectancy. The first result tells us that if the population size and structure (by age and sex) in 2019 had been the same as in 2020 (taken here as the reference year), then the number of deaths would have been 9.2% higher in 2020 than in 2019. The second result tells us that the average life span of a hypothetical cohort living and dying according to observed mortality rates in 2020 would be 0.8% (or 8 months) shorter than that of a hypothetical cohort living and dying according to observed mortality rates in 2019, the former reaching 83.1 and the latter 83.7 years (calculated over both sexes). Compared to SMR, which treats every death equally, life expectancy gives more weight to a death occurring at a young age than at an advanced age, recognizing that more years are lost in the former case. Because COVID-19 killed primarily elderly people, its impact on mortality appears less dramatic when mortality is assessed by a loss in life expectancy than by an increased SMR.

Yet, following arguments in Goldstein and Lee [7], the above loss of 8 months of life expectancy in Switzerland in 2020 attributed to COVID-19 is probably exaggerated, as it considers a hypothetical cohort that would live a life long under the mortality conditions observed in 2020. In other words, it assumes that persons in this hypothetical cohort would live their entire lives with the COVID-19 pandemic. If, as we hope, the COVID-19 pandemic lasts at most a few years (at least in its most severe form), and if the situation improves thereafter, the life lost to COVID-19 will probably amount to not a few months but a few days.

In this paper, we attempt to calculate the amount of life lost to COVID-19 in 2020 based on all-cause mortality, not for hypothetical cohorts, but for actual populations of persons living in 2020. We focus on 12 European countries for which complete mortality data were available for 2020. The calculations presented here could easily be repeated once data are available for subsequent years to assess the impact of COVID-19 on mortality across all pandemic years.

## 2. Data

We used mortality data that can be found on the Human Mortality Database (HMD [8], last accessed on April 1, 2022). This is a classic website for researchers interested in demography where one can find the remaining life expectancy as well as the number of deaths and the population size at each age between 0 and 110 years, for various countries and calendar years, separately for women and men. We selected the 12 European countries for which data were available up to the year 2020, namely Belgium (BEL), Croatia (CRO), Denmark (DEN), Finland (FIN), Hungary (HUN), Lithuania (LIT), Luxembourg (LUX), Norway (NOR), Portugal (POR), Spain (SPA), Sweden (SWE) and Switzerland (SWI). The first year for which such data were available ranged from 1751 (Sweden) to 2001 (Croatia).

## 3. Methods

Goldstein and Lee [7] considered that the population of *N*^2020^ = 330 million persons living in America in 2020 had on average a remaining life expectancy of *E*^2020^ = 45.8 years, making a total of *N*^2020^ · *E*^2020^ = 14′900 million years of remaining life for the whole population. On the other hand, they were hypothesizing (current 2020) a total of 1 million deaths due to COVID-19 among that population, with an average remaining life expectancy of 11.7 years for the deceased, making a total of *Y*^2020^ = 11.7 million years lost to COVID-19. They concluded that this loss would correspond to *Y*^2020^/(*N*^2020^ · *E*^2020^) = 11.7/14′900 = 0.08% (i.e. less than 1/1000) of the remaining life of the population living in America in 2020. If we want to express this result in terms of years, we can say that the average (i.e. per person) life lost to COVID-19 in this population, or *population life loss* for short, would be *PLL* ^2020^ = 0.08% · *E*^2020^ = *Y*^2020^/*N*^2020^ = 11.7/330 = 0.035 years, or 13 days.

In the present paper, we followed Goldstein and Lee to estimate the life lost to COVID-19 for actual populations of women and men living in 2020 in the 12 European countries above. For a given country and gender, let 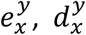 and 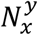 denote respectively the remaining life expectancy, the number of deaths and the size of the population at age *x* in calendar year *y*, taken from HMD. We calculated an “excess deaths’’ 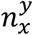 at age *x* in year *y* by comparing the number of (all-cause) deaths that year with the number of (all-cause) deaths the year before at the same age, standardized to take into account for the change in population size between the two years, yielding:

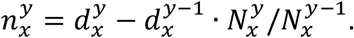

For year *y* = 2020, this quantity will be interpreted as the number of deaths at age *x* attributable directly or indirectly to COVID-19, as discussed in the Introduction. Quantities corresponding to those used by Goldstein and Lee for year *y* were then calculated as 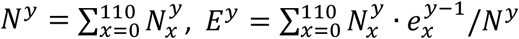 and 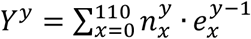. Denoting by 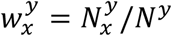 the proportion of persons of age *x* in year *y*, population life loss in year *y* is thus obtained as:

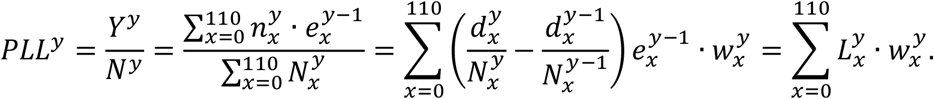

The expression on the right emphasizes that a population life loss is obtained as a weighted average of the life losses at different ages *x* in year *y* defined by:

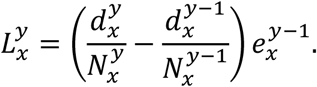

Considering that the quantities 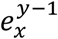 and 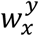 are known (fixed) for a year *y*, and that the expectations of the proportions of deaths 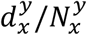 at the different ages *x* in year *y* are based on the mortality observed the year before, i.e. equal to 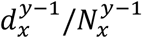, also considered as known (fixed) quantities, we calculated the following standard errors:

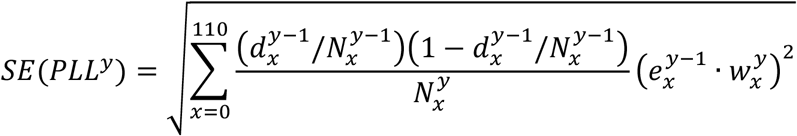

and:

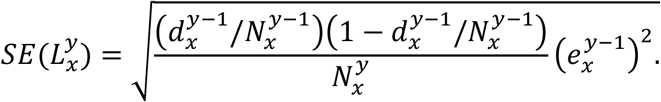

We calculated 95% confidence bands for a population life loss, respectively a life loss at age *x*, in a year *y*, under the hypothesis of a similar mortality as the year before (e.g. what would have been expected in a year *y* = 2020 without COVID-19) as ±1.96 · *SE*(*PLL*^*y*^) and 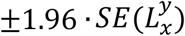. Values of *PLL*^*y*^ or 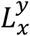 outside these bands indicated statistically significant losses.

## 4. Results

In 2019, life expectancy at birth ranged from 71.5 (Lithuania) to 81.9 (Switzerland) for men and from 79.7 (Hungary) to 86.2 (Spain) for women. By 2020, it had decreased in almost all countries, usually more for men than for women, up to 1.4 years for Lithuanian men. The exceptions were Denmark and Norway, as well as Finnish women, for whom life expectancy increased slightly in 2020 despite the pandemic. See the first three columns of Table 1 for more details. However, as mentioned in the Introduction, such a loss in life expectancy would concern a hypothetical cohort of persons living their entire lives under the mortality conditions of the COVID-19 pandemic in 2020. To quantify what the actual populations of persons living in these countries in 2020 have lost during that pandemic year, we calculated the population life loss (*PLL*^2020^) as explained in the Methods section. Results are provided in Table 1, together with the number of inhabitants (*N*^2020^), the remaining life expectancy (*E*^2020^) and the total years lost (*Y*^2020^) in these countries in 2020.

**Table 1:** Life expectancies at birth in 2019 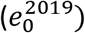 and 2020 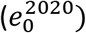, as well as life expectancy loss 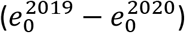, number of inhabitants (*N*^2020^), remaining life expectancy (*E*^2020^), total years lost (*Y*^2020^) and population life loss (*PLL*^2020^) in 2020 for men (M) and women (F) of 12 European countries.

| **Country** | $e_0^{2019}$<br>(years) | $e_0^{2020}$<br>(years) | $e_0^{2019} - e_0^{2020}$<br>(years) | $N^{2020}$<br>(million) | $E^{2020}$<br>(years) | $Y^{2020}$<br>(1000 years) | $PLL^{2020}$<br>(days) |
| --- | --- | --- | --- | --- | --- | --- | --- |
| **LIT M** | 71.5 | 70.1 | 1.4 | 1.3 | 35.7 | 31.2 | 8.7 |
| **LIT F** | 81.0 | 80.0 | 1.0 | 1.5 | 38.3 | 22.2 | 5.5 |
| **SPA M** | 80.8 | 79.6 | 1.3 | 23.2 | 41.1 | 320.3 | 5.0 |
| **SPA F** | 86.2 | 85.0 | 1.2 | 24.1 | 43.4 | 286.1 | 4.3 |
| **BEL M** | 79.6 | 78.5 | 1.1 | 5.7 | 41.6 | 68.4 | 4.4 |
| **BEL F** | 84.0 | 83.1 | 0.9 | 5.8 | 43.3 | 58.8 | 3.7 |
| **HUN M** | 73.0 | 72.3 | 0.7 | 4.7 | 35.9 | 51.0 | 4.0 |
| **HUN F** | 79.7 | 79.0 | 0.7 | 5.1 | 37.8 | 51.7 | 3.7 |
| **CRO M** | 75.4 | 74.7 | 0.7 | 2.0 | 36.9 | 20.0 | 3.7 |
| **CRO F** | 81.5 | 80.8 | 0.6 | 2.1 | 38.7 | 17.8 | 3.1 |
| **POR M** | 78.6 | 78.0 | 0.7 | 4.9 | 38.8 | 45.2 | 3.4 |
| **POR F** | 84.6 | 84.0 | 0.6 | 5.4 | 40.8 | 35.8 | 2.4 |
| **SWI M** | 81.9 | 81.0 | 0.9 | 4.3 | 42.8 | 35.8 | 3.1 |
| **SWI F** | 85.6 | 85.1 | 0.5 | 4.3 | 44.1 | 18.9 | 1.6 |
| **SWE M** | 81.3 | 80.6 | 0.8 | 5.2 | 43.3 | 38.7 | 2.7 |
| **SWE F** | 84.7 | 84.3 | 0.4 | 5.1 | 44.6 | 20.0 | 1.4 |
| **FIN M** | 79.2 | 79.1 | 0.1 | 2.7 | 40.4 | 2.8 | 0.4 |
| **FIN F** | 84.5 | 84.7 | -0.1 | 2.8 | 42.4 | -4.2 | -0.5 |
| **LUX M** | 80.0 | 79.8 | 0.2 | 0.3 | 43.2 | 0.2 | 0.2 |
| **LUX F** | 84.8 | 84.4 | 0.4 | 0.3 | 45.8 | 0.9 | 1.0 |
| **DEN M** | 79.4 | 79.6 | -0.1 | 2.9 | 41.1 | -6.5 | -0.8 |
| **DEN F** | 83.4 | 83.5 | -0.1 | 2.9 | 42.9 | -3.2 | -0.4 |
| **NOR M** | 81.2 | 81.5 | -0.3 | 2.7 | 43.7 | -8.5 | -1.2 |
| **NOR F** | 84.7 | 84.9 | -0.2 | 2.7 | 45.3 | -5.4 | -0.7 |

The greatest population life loss was found for Lithuanian men. As detailed in Table 1, the *N*^2020^ = 1.3 million men living in Lithuania in 2020 had on average a remaining life expectancy of *E*^2020^ = 35.7 years, making a total of 1.3 · 35.7 = 46.7 million years of remaining life for the whole population. The total years lost to COVID-19, obtained from a comparison of the all-cause mortality in 2019 and 2020, was *Y*^2020^ = 31.2 thousand years. The loss to COVID-19 for Lithuanian men therefore amounted to 31’200/46′700′000 = 0.07% of their remaining life, whereas the population life loss was *PLL* ^2020^ = 0.07% · 35.7 = 31’200/1’3000’000 = 0.024 years, or 8.7 days per person. Results of similar calculations for men and women of other countries are found in Table 1. Here and in the Figures below, countries are ordered by decreasing population life loss for men. Behind Lithuanian men, the greatest losses were observed for men in Spain (5.0 days), Belgium (4.4 days), Hungary (4.0 days), Croatia (3.7 days), Portugal (3.4 days), Switzerland (3.1 days) and Sweden (2.7 days), while women lost 5.5, 4.3, 3.7, 3.1, 2.4, 1.6 and 1.4 days, respectively, i.e. less than men in all countries. Further down the table, we find countries with negative losses, corresponding to gains, as in Denmark, Norway and for Finnish women, consistent with the gains in life expectancy mentioned above, the most important one being for Norwegian men who gained 1.2 days in 2020.

To get a comparison with other recent years, Figure 1 shows the population life loss calculated each year between 1980 and 2020 for women and men living in these 12 countries in those years. In most recent years, losses were significantly negative, indicating yearly gains of a few days, due to the steady decline in mortality along the years, and reflecting the continued progress in this domain. The consistent and significant (positive) losses of a few days observed in 8 out of the 12 countries in 2020 thus appeared to be a notable exception, illustrating the significant impact of the COVID-19 pandemic on mortality in 2020 in these countries. The four countries without a significant loss in 2020 were Finland, Luxembourg, Denmark and Norway.

**Figure 1:**
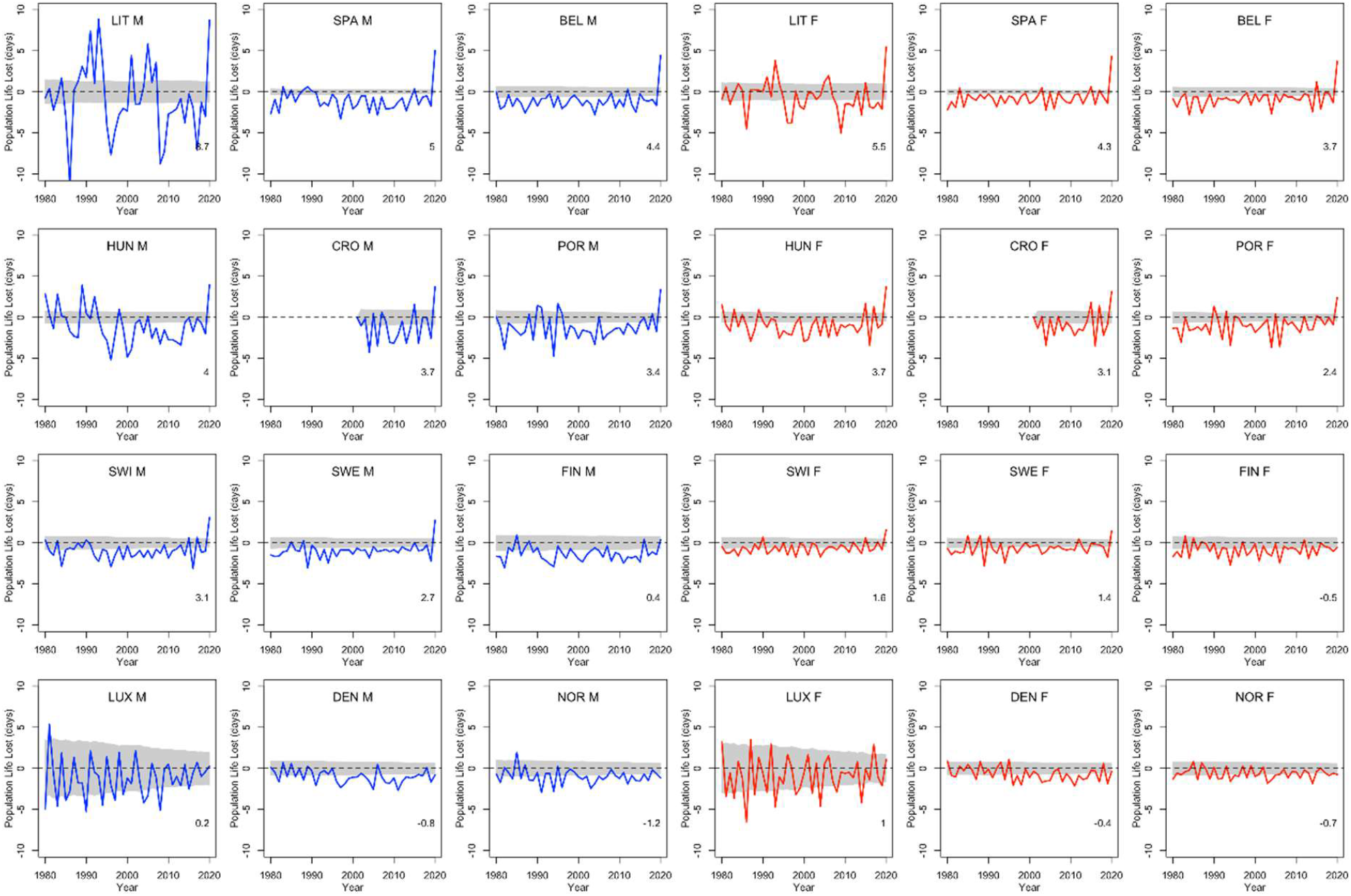
Population life loss (*PLL*^*y*^) in years *y* between 1980 and 2020 for men (M) and women (F) of 12 European countries, together with 95% confidence bands. At the bottom, population life loss in 2020 (*PLL*^2020^, expressed in days) is indicated.

To get further comparisons, we calculated the population life loss in year 1918 (compared to 1917) at the time of the Spanish flu in countries with available data. For men, the loss amounted to 10.3, 60.1, 75.5, 87.3, 135.4 and 246.5 days (per person) in Denmark, Norway, Sweden, Switzerland, Spain and Finland, respectively. For women, it was 12.0, 55.8, 65.3, 65.1, 141.6 and 42.4 days, respectively. The Spanish flu in 1918 had thus a much greater impact on mortality, on average about 30 times greater on that scale, than COVID-19 in 2020.

Figure 2 shows life losses in 2020 at the different ages between 0 and 90 (due to small sample sizes, life losses over 90 showed too much variability to get a reliable interpretation). In countries with a significant population life loss in 2020, life loss was clearly increasing with age, reaching about 40 days at the age of 90 in some countries. This trend was particularly consistent in large populations, where confidence bands were narrower, as in Spain. Based on these plots, life loss became consistently significant for men from age 47, 40, 57, 60, 68, 65, 74 and 70 years in Lithuania, Spain, Belgium, Hungary, Croatia, Portugal, Switzerland and Sweden, respectively. For women, this was the case from age 69, 52, 68, 61, 69, 69, 79 and 73 years, respectively. In contrast, no losses were consistently significant at younger ages.

**Figure 2:**
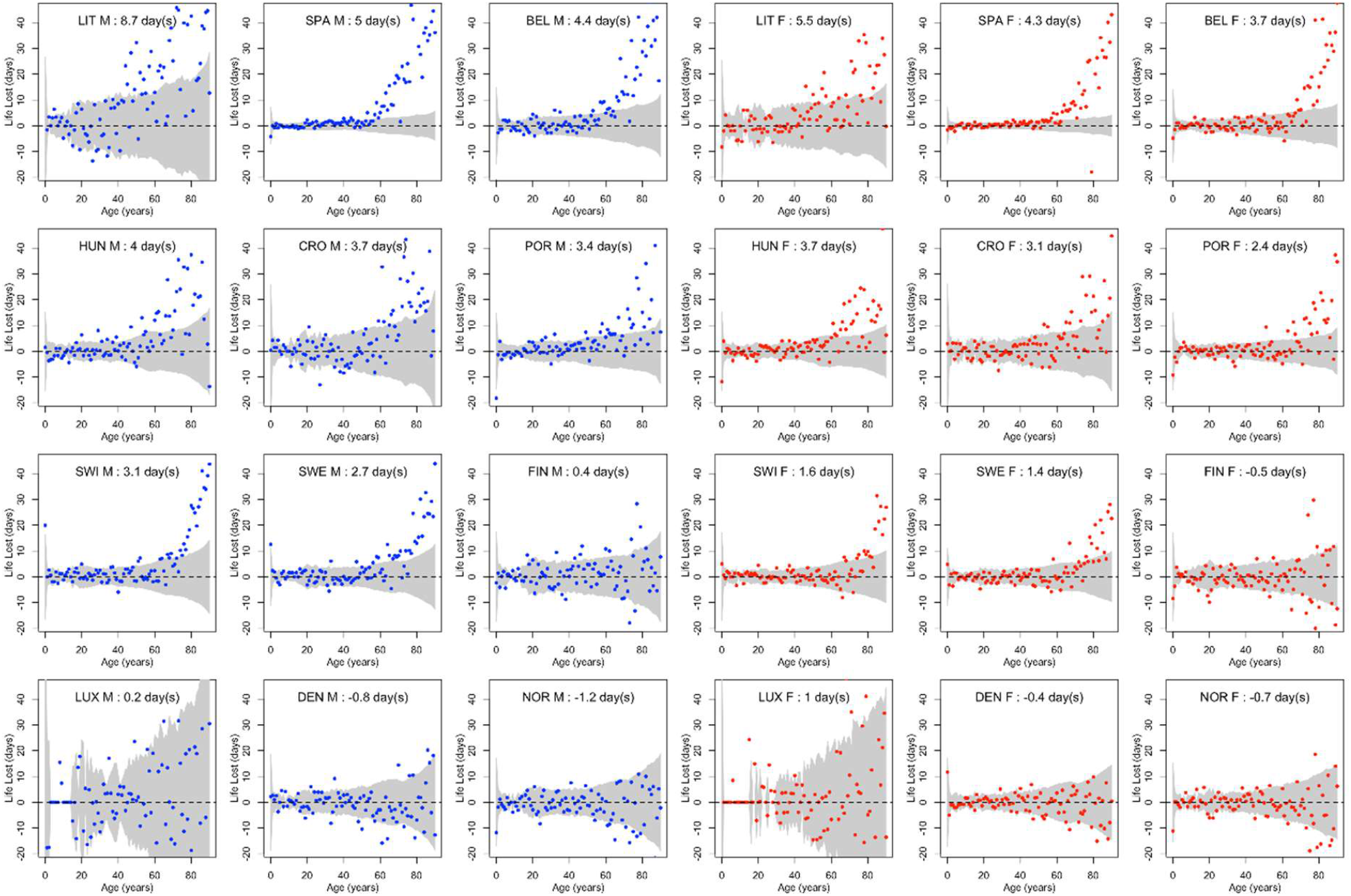
Life losses in 2020 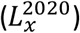 at ages *x* between 0 and 90 for men (M) and women (F) of 12 European countries, together with 95% confidence bands. At the top, population life loss in 2020 (*PLL*^2020^, expressed in days) calculated as weighted average of life losses at the different ages *x* is indicated.

## 5. Discussion

Estimating the impacts of COVID-19 on mortality has been the topic of much recent research. Many studies based their calculations on all-cause mortality, as we did. But whereas most studies used either excess deaths (calculated via standardized mortality rates, e.g. [9]) or a loss in life expectancy (e.g. [10]) to quantify the impact of COVID-19 on mortality in 2020, we used the concept of “population life lost”. As for life expectancy, this gives more weight to a death at a young age than to a death at an advanced age. But in contrast to life expectancy, it considers the life lost for an actual population of persons living during a pandemic year, not for a hypothetical cohort of persons who would live their entire lives with such a pandemic. Using that concept, we could retrieve well-established results, e.g. that the COVID-19 pandemic affected more men than women, and mostly the elderly, while some countries (like Lithuania, Spain or Belgium) were more affected than others (like Finland, Denmark or Norway), as found e.g. in [9-10]. We could also confirm that the Spanish flu of 1918 had a much greater impact on mortality than COVID-19 in 2020 [11]. But while life expectancy losses in 2020 amounted to a few months (or a little more than one year) in most of the countries considered, population life losses in 2020 amounted to a few days, so the impacts of the COVID-19 pandemic on the mortality may appear less dramatic on that scale.

Other studies calculated a number of years of potential life lost to COVID-19, such as [12], where a total of 20.5 million years were counted in 2020 (or in fact up to January 6, 2021) across 81 countries affected by COVID-19 worldwide. This quantity was calculated as the sum of the remaining life expectancies at the time of death over all individual deaths attributed to COVID-19, following the method suggested by Greville [13]. Such a total is sometimes divided by the number of deaths, or by a number of person-years [14-15]. One serious interpretational issue with this concept, however, is that it is always positive by definition, since a remaining life expectancy is necessarily larger than zero, even at an advanced age. As a consequence, it cannot be naturally compared with zero and it is not obvious to get a sensible reference value to judge of the importance of a given amount of years lost [14]. This is why this concept is mostly used in a relative sense for comparison purposes, e.g. to compare the burden of different diseases as in Global Burden of Disease Studies [16], rather than being interpreted at face value, i.e. in an absolute sense [17-18].

By way of contrast, the concept of population life lost, *PLL*^*y*^ = *Y*^*y*^/*N*^*y*^, as implemented here, can take on negative values, due to the possibility of our numerator to be negative, as was the case in Denmark, Norway and for Finnish women in 2020, and in all countries in most pre-pandemic years. Consequently, the zero value is attainable and is thus a natural reference value. While we still interpret our numerator *Y*^*y*^ as a total number of years lost to COVID-19 in year *y* = 2020, it is not based on the deaths specifically attributed to COVID-19, but is obtained via a comparison of all-cause mortality with the year before. It will thus be negative (respectively equal to zero) if the mortality in year *y* is found to be lower than (respectively equal to) the mortality the year before, and positive otherwise. We then used the population size *N*^*y*^ as denominator to get a “population measure”, as the population is the primary object of interest in public health.

One matter of discussion is that various restrictive lockdown (among other) policies have been implemented in most countries in 2020 and it is difficult to guess what mortality would have been without these measures [19]. A notable exception is Sweden, where only soft measures have been taken [20]. Since COVID-19 mortality was higher than in neighboring (comparable) countries, Sweden has been criticized in this regard [21-22]. It is thus interesting to mention that the population life loss in 2020 over both sexes was 2.1 days per person in Sweden, whereas it was -0.1, -0.7 and -0.9 days (corresponding to gains) in Finland, Denmark and Norway, respectively. If we deduce from there that Sweden would have gained up to 0.9 days (instead of losing 2.1 days) by applying a lockdown similar to that of neighboring countries, the cost of not having done any lockdown at the level of the Swedish population (i.e. in a public health perspective) might be estimated at 2.1+0.9=3 days of life per person. One (inevitably controversial) question here is how many days of lockdown would be acceptable for a population to save up to 3 days of life? Of course, this is a highly theoretical and necessarily speculative question but these kinds of calculations are considered in other contexts and domains, e.g. to establish the maximum cost of a drug that is acceptable to save one year of life [23-24].

While the proposed concept of population life loss can be readily implemented from conventional official and demographic statistics, it also suffers from technical limitations, which could be improved with more sophisticated models. One is that we are comparing the observed mortality a given year with that of the year before to calculate our numerator. To calculate the number of years lost to COVID-19 in 2020, we compared the mortality in 2020 with that of 2019, implicitly assuming that mortality would have been the same in 2020 as in 2019 without COVID-19. We have therefore ignored the secular yearly decline in mortality, while making all our calculations dependent on a single year, with the risk that this year might be a special (outlying) one. Modeling the mortality decline in recent years would improve these points, although the result may depend significantly on the model chosen and the number of recent years considered. Using mortality data from 2010-2019, we tentatively estimated a yearly mortality decline of about 2% in Switzerland. Assuming and accounting for such a decline in a year 2020 without COVID-19, our estimates of population life losses in 2020 would be increased by about 20%-30% compared to those provided in Table 1, becoming e.g. 3.6 and 2.1 (instead of 3.1 and 1.6) days for Swiss men and women, respectively. So our conclusions would not be drastically different.

Another point is that the remaining life expectancies at each age are taken from period life tables, not from cohort life tables, which are not available for recent years while not obvious to calculate [25], again implicitly assuming that mortality would remain stable in a future without COVID-19. But if mortality continues to decline, remaining life expectancies from period life tables will underestimate reality, may be by 10%. For example, scenarios (inspired by real data) for simulations in Sweden considered a period life expectancy rising from 47 to 95 years between 1850 and 2150, with corresponding cohort life expectancies rising from 51 to 102 years, i.e. close to (although less than) 10% higher [26]. This underestimation might be partially offset, however, by the fact that those who are dying at an age *x* may not be fully representative of the entire population at that age, perhaps being more frail than average, and thus with a lower remaining life expectancy, for example because of multimorbidity [15].

As lost days can be sensibly added up to lost days, we intend to repeat such a calculation for 2021 and subsequent years, and add up the lost days calculated across all pandemic years. This will be only relevant, though, if no other major events that could affect mortality (such as wars or economic recessions) occur during those years. Otherwise, the impact of COVID-19 on mortality so calculated will be confounded by the impacts of these events.

We conclude with a quotation of Goldstein and Lee [7], from which the present article is inspired, who wrote about COVID-19 in America that “it is possible to portray the epidemic as unimaginably large - the biggest killer in American history - or small, reducing our remaining life by less than 1 part in 1000”. Using the concept of population life loss, we were able to confirm the significant impact of COVID-19 on mortality in 8 out of 12 European countries in 2020. We could also show that the life lost to COVID-19 for actual populations living in 2020 in these countries can be counted in days rather than months or years, a potentially useful piece of information from a public health perspective.

## Data Availability

All results calculated in the present study are based on data freely available at www.mortality.org

